# Evaluation of Long-Term Amyotrophic Lateral Sclerosis Survivors Treated with Masitinib in Study AB10015

**DOI:** 10.64898/2026.04.10.26350104

**Authors:** Albert C. Ludolph, Terry Heiman-Patterson, Jesus S. Mora, Javier Mascías Cadavid, Gabriel Rodriguez, Natalia Bohorquez Morera, Patrick Vermersch, Alain Moussy, Colin D Mansfield, Olivier Hermine

## Abstract

**Introduction:** Amyotrophic lateral sclerosis (ALS) is a progressive neurodegenerative disease with limited treatment options. Masitinib, a tyrosine kinase inhibitor targeting microglial and mast cell activity in ALS pathogenesis, offers potential neuroprotection. This study presents a post-hoc analysis of long-term survivors treated with masitinib at 4.5 mg/kg/day in study AB10015, comparing observed survival to predicted and historical benchmarks.

**Methods:** Study AB10015 was a randomized, double-blind, placebo-controlled trial assessing masitinib with riluzole in ALS patients. Overall survival (OS) was measured from symptom onset to death, encompassing the double-blind period and post-study follow-up, including an optional, open-label program. The ENCALS model predicted survival of long-term survivors (≥5 years). A delay in the need for mechanical assistance, such as permanent ventilation, gastrostomy, tracheostomy, or wheelchair dependence, was used as a surrogate measure for quality of life (QoL).

**Results:** Among 130 patients receiving masitinib 4.5 mg/kg/day, the 5-year survival rate from onset was 42.3%, increasing to 50.0% in patients with an ALSFRS-R progression rate from disease onset of <1.1 points/month (AB10015 primary efficacy population) and 52.9% in a subgroup of patients without complete loss of functionality at baseline. Half of the long-term survivors had satisfactory QoL, defined as no mechanical assistance. The median OS for long-term survivors (n=55) was 121 months versus the ENCALS-predicted 42 months, yielding a 79-month residual median survival gain. Long-term survivors were prevalent across ALS baseline prognostic factors, including slow or moderate disease progression rate (ΔFS), severe or moderate functional severity, bulbar or spinal site of onset, respiratory function and age. Long-term survival was less likely in patients with complete loss of function at baseline or fast progressing disease (ΔFS ≥1.1 points/month) at baseline.

**Conclusions:** Masitinib treatment in ALS patients showed substantial survival benefit. Long-term survivors were largely independent of ALS prognostic factors, suggesting a subpopulation driven by microglial/mast cell activity. A recently identified biomarker detecting masitinib’s effect on pro-inflammatory microglia may help identify responsive patients.

## Introduction

Amyotrophic lateral sclerosis (ALS) is a debilitating and life-threatening disease characterized by progressive loss of movement, respiratory insufficiency, and poor survival rates. No treatment has been proven to meaningfully enhance survival. The ongoing challenges in developing broadly effective treatments for ALS are evident in the modest survival benefits associated with riluzole, the standard first-line drug for ALS for the past three decades [Miller 2012], inconsistencies in post-marketing efficacy data for edaravone [Gupta 2025, Kashyap 2025, Takahashi 2025, Shefner 2024, Witzel 2022, Brooks 2022, Ortiz 2020, Lunetta 2020, Fortuna 2019], and a shift towards more targeted therapies, such as tofersen, for rare genetic (familial) subtypes of ALS [Miller 2022]. Consequently, there is an urgent need for novel therapies that can delay disease progression in patients with ALS.

Masitinib is a selective tyrosine kinase inhibitor that exerts neuroprotection in both central and peripheral nervous systems via selective kinase inhibition that modulates the functionality of different cells implicated in ALS pathogenesis. By targeting the CSF1/CSF-1R signaling pathway, masitinib has the potential to regulate CSF-1R-dependent cells, such as microglia, which are implicated in ALS progression [Brites 2014, Clarke 2020]. Additionally, its actions against KIT, LYN, and FYN allow masitinib to inhibit mast cell function and activation, which are crucial effector immune cells in chronic inflammatory processes [Krystel-Whittemore 2016, Angelini 2020] and interact with microglia and astrocytes in ALS, potentially leading to motor neuron damage [Skaper 2018, Vahsen 2021, Sandhu 2021].

Cohort- and population-based studies indicate that the median survival from symptom onset to death for ALS ranges from 26 to 44 months (approximately 2.2 to 3.7 years), varying by region, cohort type, and the use of interventions, such as ventilation or tracheostomy [Pupillo 2025; Engelberg-Cook 2024; Chio 2023; Spittel 2021; Calvo 2017]. Although there is no standard definition of ‘long survival’ in ALS, the literature indicates that surviving at least 5 years from the onset of symptoms surpasses expectations regarding median survival and its associated upper-quartile boundary. This is also reflected in the low benchmark 5-year survival probability of approximately 23.5% (Table 1) [Pupillo 2025; Bianchi 2022; Jennum 2013; Mateen 2010; del Aguila 2003; Preux 1996; Christensen 1990]. Therefore, long-term ALS survivors are defined here as patients living for at least 5 years after disease onset.

**Table 1:** Studies reporting 5-Year survival in ALS.

| Study ID | N | 5-year survival rate | Comment |
| --- | --- | --- | --- |
| Pupillo 2025 | 828 | 21.0% <sup>2</sup> | Italian population-based registry (SLALOM) |
| Bianchi 2022 | 267 | 24.0% <sup>2</sup> | Italian population-based registry (SLALOM) |
| Jennum 2013 | 2,394 | 27.8% | Danish national patient registry |
| Mateen 2010 | 94 | 14.0% <sup>1</sup> | Olmsted County, Minnesota, USA |
| del Aguila 2003 | 180 | 7.0% <sup>2</sup> | King County, Washington, USA |
| Preux 1996 | 158 | 14.7% | French cohort |
| Christensen 1990 | 186 | 7.0% <sup>1</sup> | 2 Danish counties |
| **Weighted Average** |  | **23.5%** |  |

<sup>1</sup> 5-year survival rate from the symptom onset. <sup>2</sup> 5-year survival rate from diagnosis.

This article details the results of a post-hoc analysis of patients from study AB10015 (NCT02588677) [Mora 2020] and its associated early access program, concentrating on the 4.5 mg/kg/day treatment arm, as this dose was selected for the future development of masitinib in ALS. It compares observed survival from disease onset with a benchmark 5-year survival rate from the literature and also with predicted survival using the European Network for the Cure of ALS (ENCALS) personalized survival model for a cohort of long-term survivors (i.e., AB10015 patients surviving ≥5 years from disease symptom onset). The aim was, in part, to provide a better context for reports of notable longevity from sites participating in the early access program of masitinib.

Study AB10015 was an international, phase 2b/3, randomized, placebo-controlled trial that evaluated oral masitinib as an add-on therapy to riluzole (100 mg/day) in ALS [Mora 2020]. The primary endpoint was the change in the revised amyotrophic lateral sclerosis functional rating scale (ΔALSFRS-R) after 48 weeks of treatment. The prespecified primary efficacy analysis population included patients with an ALSFRS-R progression rate from disease onset to baseline (ΔFS) of less than 1.1 points/month and who received masitinib at 4.5 mg/kg/day. The results for masitinib 4.5 mg/kg/day indicated an acceptable safety profile in patients with ALS and a significant benefit over placebo in ΔALSFRS-R, with a 3.39-point difference (p=0.016), representing a clinically meaningful 27% decrease in ALSFRS-R decline over 48 weeks compared with placebo treatment. This finding was confirmed by multiple sensitivity analyses [Mora 2020]. Additionally, results from an overall survival (OS) analysis indicated that masitinib 4.5 mg/kg/day could prolong survival relative to placebo by +6 months in the primary analysis population (47 months (95%CI [30; 69]) versus 41 months (95%CI [30; 54]), respectively) [Mora 2021], and by a statistically significant +12 months relative to placebo (p=0.0192) in the subgroup ‘ALS prior to any complete loss of functionality’ (53 months (95%CI [36; NE]) versus 41 months (95%CI [30; 54]), respectively) [Ludolph 2025].

## Methods

Details regarding the method and patient selection criteria of study AB10015 have been previously published [Mora 2020]. Briefly, patient randomization was conducted using a minimization method based on baseline covariates, including the site of onset (spinal versus bulbar), ALSFRS-R score, age, geographical region, and ΔFS. Patients receiving masitinib 4.5 mg/kg/day with a post-onset ΔFS of less than 1.1 points/month were designated as the primary efficacy analysis population. Eligible patients were aged 18–75 years with a laboratory-supported probable, probable, or definite diagnosis of ALS, a disease duration of less than 36 months from the first ALS symptom and forced vital capacity (FVC) of at least 60% at baseline. Additionally, patients were required to be on a stable dose of riluzole (100 mg/day) for at least 30 days prior to baseline. Following data readout for the blinded protocol period of study AB10015, the treatment assignment was unblinded, and an optional open-label early access program was initiated. This allowed patients who were still receiving masitinib to continue treatment, with all patients receiving the recommended masitinib dose of 4.5 mg/kg/day. The protocol for study AB10015 was approved by the institutional review board or ethics committee at each of the 34 participating clinical site (including the Secretaría Técnica del Comité Ético de Investigación Clínica, Clínica del Hospital Universitario La Paz, Madrid, Spain) and conducted in accordance with the Declaration of Helsinki. All patients provided written informed consent, which included the provision to review medical records for the completion of undetermined endpoints, such as overall survival.

Overall survival for participants in study AB10015 was defined as the time elapsed between symptom onset and death from any cause. Kaplan–Meier estimates were used to calculate the survival times. Patients whose endpoint was not death were censored on the date of their last follow-up contact. Overall survival p-values were calculated via the log-rank test

The ENCALS survival prediction model, based on data from over 11,000 patients with ALS in population-based registers, was developed to predict personalized survival for patients with ALS, with outcomes reported in terms of prognostic groups or as a point estimate within a survival curve. The development of the ENCALS survival prediction model has been previously described [Westeneng 2018]. Briefly, it combines multiple clinical and biological factors known to be associated with ALS outcomes into a multivariate survival model to predict a composite survival outcome (time between the onset of symptoms and noninvasive ventilation >23 h/day, tracheostomy, or death) in individual patients. We used the ENCALS survival prediction model to retrospectively calculate the expected survival of long-term survivors from the masitinib 4.5 mg/kg/day treatment arm of study AB10015. Multiple clinical and biological factors were collected from patient records and entered into the ENCALS survival prediction tool (http://tool.encalssurvivalmodel.org/). These included age at onset, diagnostic delay, site of symptom onset (limb vs. bulbar), diagnostic classification of definite ALS (according to the El Escorial criteria), ALSFRS-R slope, and lung function (forced vital capacity percentage). Patients were ineligible for study AB10015 if they presented with dementia [Mora 2020]; hence, the presence of frontotemporal dementia (cognitive factor) was entered as negative for all patients. Data pertaining to the presence of the C9orf72 mutation were unavailable; therefore, all patients were considered to be negative. The predicted survival time since onset was determined by identifying the median (0.5) survival probability on the resultant personalized survival curve, where the x-axis represents time since onset in months and the y-axis indicates survival probability ranging from 0 to 1.0.

For neurodegenerative diseases like ALS, the therapeutic goal is to increase survival while preserving a satisfactory quality of life (QoL). No formal assessment of QoL, such as patient-reported questionnaires, was conducted over the timeframe of this long-term follow-up; therefore, a surrogate measure of QoL in long-term survivors was defined as the absence of permanent ventilation, gastrostomy, tracheostomy, or wheelchair dependence.

## Results

### Participant follow-up

The long-term assessment of patients treated with masitinib in study AB10015 encompassed a prospectively declared 48-week treatment period with an associated double-blind extension (commencing in April 2013 until data readout) and a post-study (unblinded) follow-up period that included an optional compassionate use program. After the end of the study, only patients who received masitinib through the compassionate use program were followed, with a cutoff date of July 2024.

### Five-year survival rate comparison to benchmark

The 5-year survival rate from onset of symptoms in the masitinib 4.5 mg/kg/day group was 42.3% (55/130) (Table 2). The 5-year survival rate from the onset of symptoms increased to 50.0% (53/106) for patients with baseline ΔFS of less than 1.1 points/month (i.e., the designated primary efficacy analysis population of study AB10015) and increased further to 52.9% (45/85) in a subgroup of this cohort defined by patients with a score of at least 1 on all ALSFRS-R items at baseline (i.e., ALS prior to any complete loss of functionality).

**Table 2:** Observed and predicted survival of individual long-term ALS survivors^1^ from the masitinib 4.5 mg/kg/day treatment-arm of study AB10015 (n = 55)

| Patient baseline characteristics |  |  |  |  |  |  | Survival statistics (mo) |  |  |  |  |
| --- | --- | --- | --- | --- | --- | --- | --- | --- | --- | --- | --- |
| Patient ID | Onset site | ALS Severity<sup>2</sup> | ΔFS<sup>3</sup> | ALSFRS-R | Definite ALS<sup>4</sup> | FVC | Observed | Predicted<sup>5</sup> | Residual<sup>6</sup> | Death<sup>7</sup> | QoL<sup>8</sup> |
| Patient #1 | Spinal | Moderate | Slow | 43 | Yes | 99 | 78.85 | 62 | 16.85 | N | Y |
| Patient #2 | Spinal | Severe | Moderate | 32 | Yes | 78 | 80.49 | 45 | 35.49 | N | N |
| Patient #3* | Spinal | Severe | Moderate | 39 | Yes | 92 | 107.63 | 42 | 65.63 | N | N |
| Patient #4* | Bulbar | Very Severe | Moderate | 36 | Yes | 69 | 116.93 | 32 | 84.93 | N | N |
| Patient #5 | Spinal | Moderate | Slow | 46 | No | 122 | 87.63 | 69 | 18.63 | Y | N |
| Patient #6 | Bulbar | Moderate | Moderate | 45 | No | 126 | 74.68 | 28 | 46.68 | Y | N |
| Patient #7* | Spinal | Severe | Moderate | 36 | No | 104 | 114.7 | 50 | 64.7 | N | N |
| Patient #8 | Spinal | Moderate | Slow | 46 | No | 106 | 86.37 | 84 | 2.37 | N | Y |
| Patient #9 | Spinal | Very Severe | Fast | 31 | No | 71 | 66.07 | 20 | 46.07 | N | Y |
| Patient #10 | Bulbar | Moderate | Moderate | 37 | Yes | 60 | 97.11 | 24 | 73.11 | N | N |
| Patient #11 | Spinal | Severe | Slow | 41 | No | 95 | 87.88 | 70 | 17.88 | Y | N |
| Patient #12* | Spinal | Moderate | Moderate | 43 | No | 99 | 118.08 | 82 | 36.08 | N | Y |
| Patient #13* | Spinal | Severe | Moderate | 38 | No | 99 | 138.05 | 70 | 68.05 | N | Y |
| Patient #14 | Bulbar | Severe | Moderate | 39 | Yes | 91 | 73.13 | 27 | 46.13 | N | N |
| Patient #15 | Spinal | Moderate | Fast | 38 | No | 71 | 73.59 | 17 | 56.59 | N | N |
| Patient #16 | Spinal | Moderate | Slow | 41 | Yes | 73 | 62.58 | 56 | 6.58 | Y | N |
| Patient #17 | Spinal | Moderate | Moderate | 44 | No | 101 | 73.96 | 25 | 48.96 | N | Y |
| Patient #18* | Bulbar | Severe | Moderate | 37 | No | 102 | 148.08 | 50 | 98.08 | N | Y |
| Patient #19 | Bulbar | Very Severe | Moderate | 30 | No | 80 | 67.58 | 35 | 32.58 | Y | Y |
| Patient #20 | Bulbar | Severe | Moderate | 32 | No | 78 | 79.74 | 46 | 33.74 | N | N |
| Patient #21 | Spinal | Severe | Moderate | 34 | No | 64 | 76.39 | 32 | 44.39 | N | N |
| Patient #22 | Spinal | Very Severe | Moderate | 34 | No | 67 | 72.41 | 27 | 45.41 | Y | N |
| Patient #23 | Spinal | Moderate | Slow | 44 | Yes | 99 | 94.03 | 73 | 21.03 | N | Y |
| Patient #24 | Spinal | Moderate | Moderate | 37 | No | 95 | 90.06 | 70 | 20.06 | N | Y |
| Patient #25 | Bulbar | Moderate | Moderate | 41 | No | 71 | 73.5 | 33 | 40.5 | N | Y |
| Patient #26 | Spinal | Very Severe | Moderate | 34 | No | 94 | 87.19 | 40 | 47.19 | N | N |
| Patient #27 | Spinal | Severe | Moderate | 35 | No | 126 | 78.02 | 47 | 31.02 | Y | N |
| Patient #28 | Spinal | Severe | Moderate | 38 | No | 95 | 75.5 | 33 | 42.5 | N | N |
| Patient #29 | Spinal | Moderate | Moderate | 42 | No | 98 | 63.05 | 29 | 34.05 | N | Y |
| Patient #30 | Spinal | Moderate | Slow | 44 | No | 104 | 78.13 | 77 | 1.13 | Y | Y |
| Patient #31 | Spinal | Very Severe | Moderate | 23 | No | 63 | 85.42 | 27 | 58.42 | N | N |
| Patient #32 | Spinal | Severe | Moderate | 33 | No | 67 | 71.85 | 36 | 35.85 | N | N |
| Patient #33 | Spinal | Severe | Moderate | 34 | Yes | 76 | 90.81 | 29 | 61.81 | N | Y |
| Patient #34 | Spinal | Moderate | Slow | 43 | Yes | 85 | 94.33 | 44 | 50.33 | N | N |
| Patient #35 | Spinal | Moderate | Moderate | 39 | Yes | 81 | 91.66 | 38 | 53.66 | N | Y |
| Patient #36* | Spinal | Moderate | Slow | 47 | Yes | 90 | 120.74 | 81 | 39.74 | Y | Y |
| Patient ID | Onset site | ALS Severity<sup>2</sup> | ΔFS<sup>3</sup> | ALSFRS-R | Definite ALS<sup>4</sup> | FVC | Observed | Predicted<sup>5</sup> | Residual<sup>6</sup> | Death<sup>7</sup> | QoL<sup>8</sup> |
| Patient #37 | Spinal | Severe | Moderate | 34 | Yes | 88 | 95.81 | 39 | 56.81 | N | N |
| Patient #38 | Spinal | Severe | Moderate | 34 | Yes | 95 | 108.25 | 41 | 67.25 | Y | N |
| Patient #39* | Spinal | Moderate | Moderate | 38 | Yes | 96 | 111.35 | 66 | 45.35 | N | Y |
| Patient #40 | Spinal | Severe | Slow | 41 | Yes | 95 | 64.98 | 51 | 13.98 | Y | Y |
| Patient #41 | Spinal | Moderate | Slow | 42 | Yes | 91 | 84.24 | 37 | 47.24 | N | Y |
| Patient #42 | Spinal | Moderate | Moderate | 42 | Yes | 60 | 75.07 | 30 | 45.07 | Y | N |
| Patient #43 | Spinal | Moderate | Moderate | 45 | Yes | 118 | 77.83 | 30 | 47.83 | N | Y |
| Patient #44 | Spinal | Severe | Moderate | 43 | Yes | 103 | 84.24 | 47 | 37.24 | N | N |
| Patient #45* | Spinal | Moderate | Slow | 42 | Yes | 104 | 120.48 | 41 | 79.48 | N | Y |
| Patient #46 | Spinal | Very Severe | Moderate | 25 | Yes | 60 | 98.7 | 29 | 69.7 | N | N |
| Patient #47 | Spinal | Moderate | Slow | 46 | Yes | 96 | 64.13 | 43 | 21.13 | N | Y |
| Patient #48 | Spinal | Moderate | Moderate | 45 | No | 100 | 77.08 | 40 | 37.08 | Y | Y |
| Patient #49 | Bulbar | Moderate | Slow | 47 | No | 96 | 97.38 | 115 | -17.62 | Y | Y |
| Patient #50 | Spinal | Very Severe | Moderate | 31 | Yes | 94 | 93.08 | 28 | 65.08 | N | N |
| Patient #51 | Spinal | Very Severe | Moderate | 33 | Yes | 63 | 62.52 | 43 | 19.52 | Y | N |
| Patient #52 | Spinal | Severe | Moderate | 33 | Yes | 97 | 86.74 | 59 | 27.74 | N | Y |
| Patient #53 | Spinal | Moderate | Slow | 42 | No | 97 | 76.74 | 84 | -7.26 | N | N |
| Patient #54 | Spinal | Moderate | Slow | 46 | Yes | 70 | 64.2 | 45 | 19.2 | Y | Y |
| Patient #55 | Spinal | Moderate | Slow | 43 | Yes | 101 | 84.34 | 66 | 18.34 | N | Y |

The corresponding 5-year survival rates from the time of diagnosis were 36.2% (47/130) in the masitinib 4.5 mg/kg/day group, 42.9% (45/106) in patients with baseline ΔFS of less than 1.1 points/month, and 44.7% (38/85) in the subgroup ALS prior to any complete loss of functionality.

### Delay in need of mechanical assistance (quality of life surrogate measure)

Among the long-term survivors treated with masitinib at 4.5 mg/kg/day, 49.1% (27/55) maintained a satisfactory QoL, defined as the absence of the need for mechanical assistance, such as permanent ventilation, gastrostomy, tracheostomy, or wheelchair dependence. In the primary efficacy analysis population with a baseline ΔFS of less than 1.1 points/month, approximately half (49.1%, 26/53) also maintained a satisfactory QoL. This proportion increased to 55.6% (25/45) in a subgroup of patients with ALS prior to any complete loss of functionality (i.e., a score of at least 1 on all ALSFRS-R items at baseline). These findings indicate that functional independence despite disease progression was retained by a substantial subset of long-term survivors.

In the treatment arm receiving masitinib at 4.5 mg/kg/day, 20.8% (27/130) of patients were able to maintain a satisfactory QoL.

### Comparison of observed and predicted survival

Table 2 presents the observed and predicted survival of long-term ALS survivors from the masitinib 4.5 mg/kg/day treatment arm of study AB10015. In this cohort, 1.8% (1/55) had survival less than predicted, 5.5% (3/55) matched the predicted survival (i.e., within ±10%), and 92.7% (51/55) had survival greater than predicted.

Based on the Kaplan-Meier analysis (cutoff date of July 2024), the median overall survival (OS) observed from the onset of symptoms was 121 months (95% CI [97 – NR]), with 39 patients (70.9%) censored as they were still alive. In contrast, the ENCALS model predicted a median OS of 42 months (95% CI [36 – 44]) for this group, with no patients expected to survive (Figure 1). This resulted in a statistically significant residual median survival gain of 79 months (p <0.0001).

**Figure 1.**
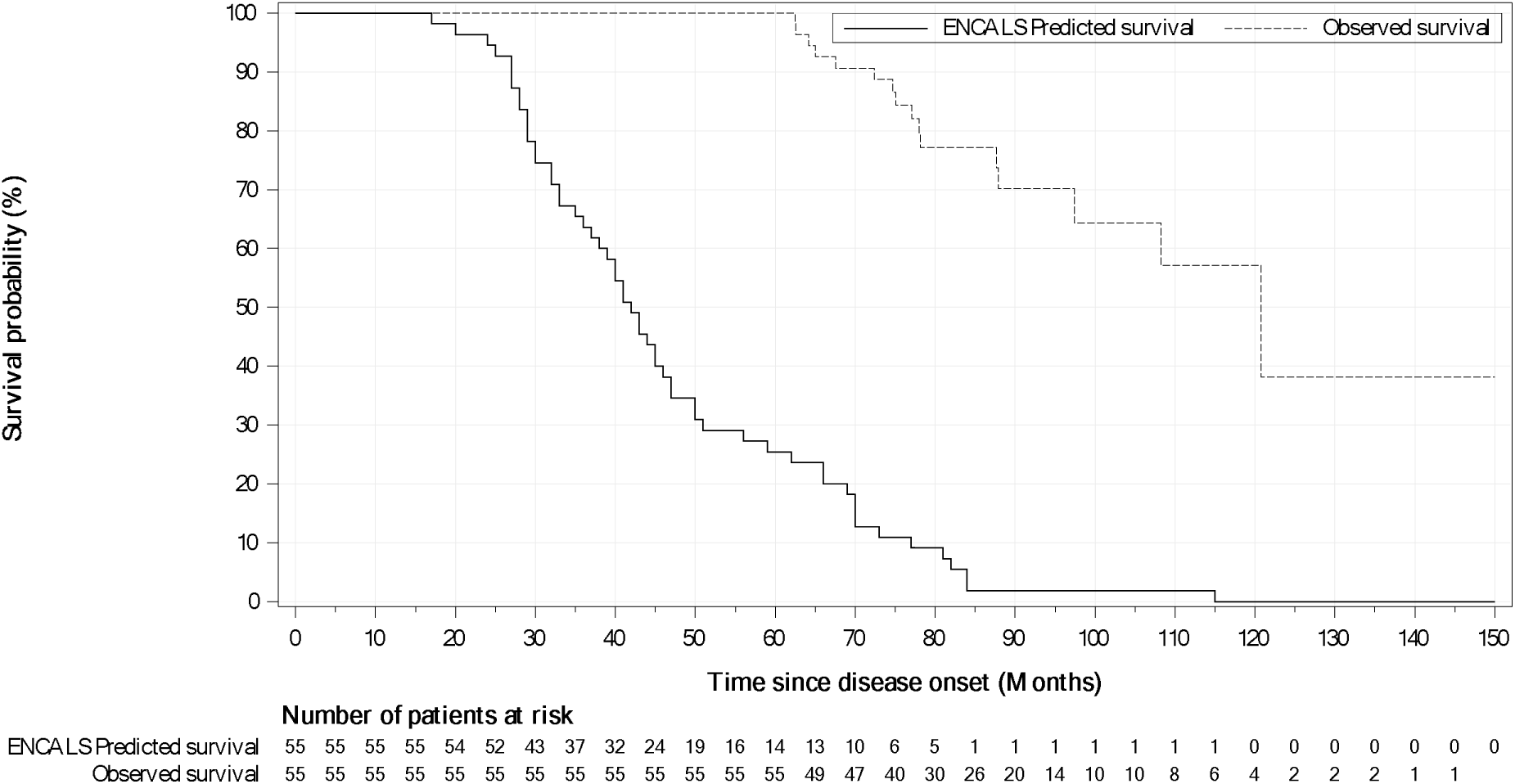
Kaplan-Meier analysis of observed survival versus ENCALS predicted survival for long-term ALS survivors^1^ treated with masitinib (4.5 mg/kg/day) (n=55)

Similarly, the observed mean average OS was calculated to be 87.3 months (SD ±19.1), compared to 47.0 months (SD ±20.3) for the ENCALS model, resulting in a 40-month (SD ±23.2) survival benefit (Table 2).

An assessment of the factors associated with masitinib-treated long-term survival in this cohort is presented in Table 3, which compares baseline predictive factors between long-term ALS survivors from the masitinib 4.5 mg/kg/day cohort and the overall masitinib 4.5 mg/kg/day treatment arm. This evaluation aimed to identify predictive factors showing a distinct shift in prevalence or magnitude in the long-term survivor group compared to the broader cohort, suggesting their potential role in treatment response. Factors with neutral (≤ 20% prevalence change in long-term survivors) or positive (more prevalent in long-term survivors) relative changes may indicate better survival outcomes, whereas those with a negative relative change (less prevalent) may be linked to poorer prognosis or a reduced likelihood of long-term survival.

**Table 3:**
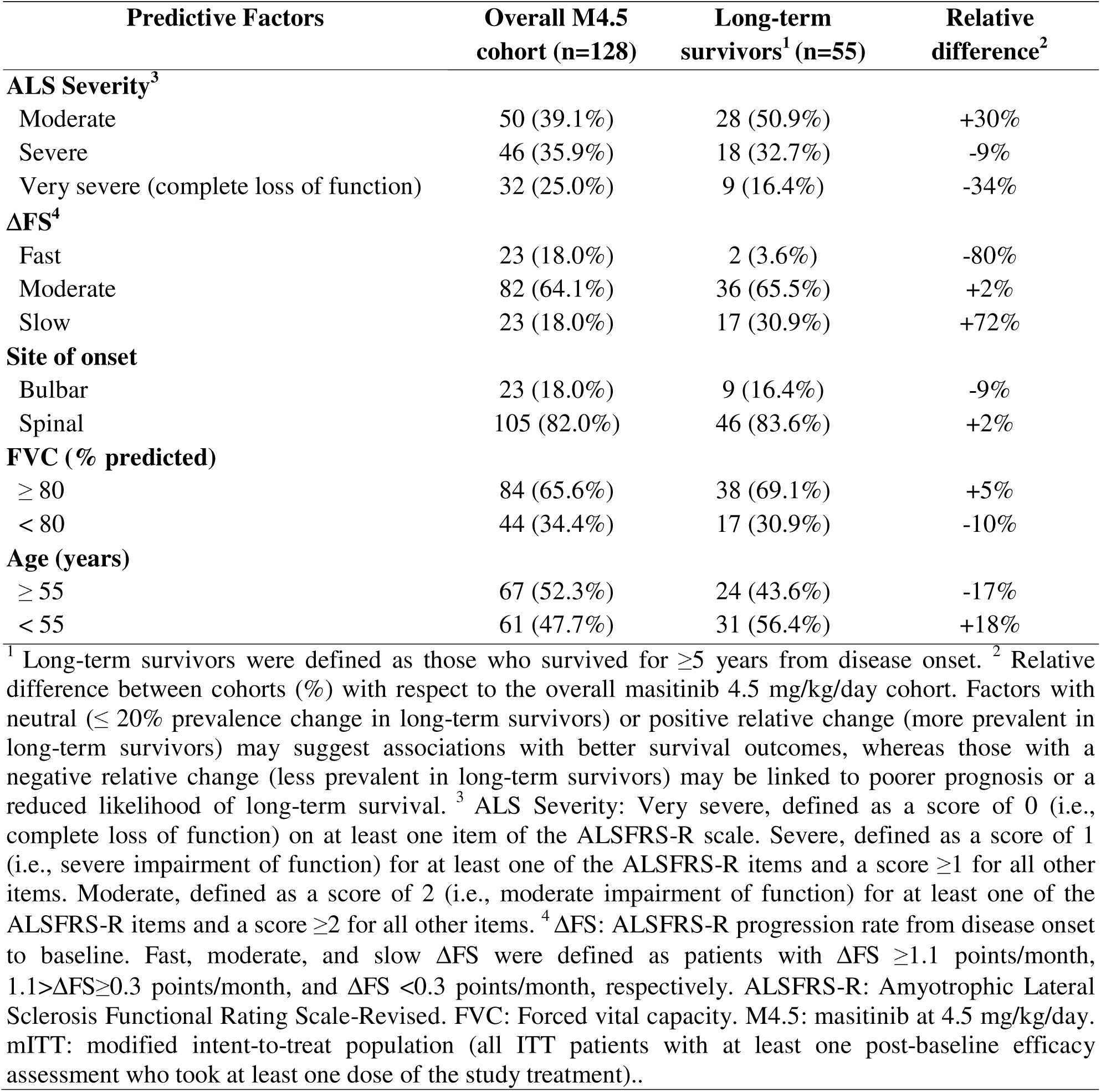
Relative change in distribution of baseline predictive factors between masitinib 4.5 mg/kg/day long-term ALS survivors^1^ and the overall masitinib 4.5 mg/kg/day cohort (mITT)

| Predictive Factors | Overall M4.5 cohort (n=128) | Long-term survivors<sup>1</sup> (n=55) | Relative difference<sup>2</sup> |
| --- | --- | --- | --- |
| **ALS Severity<sup>3</sup>** |  |  |  |
| Moderate | 50 (39.1%) | 28 (50.9%) | +30% |
| Severe | 46 (35.9%) | 18 (32.7%) | -9% |
| Very severe (complete loss of function) | 32 (25.0%) | 9 (16.4%) | -34% |
| **ΔFS<sup>4</sup>** |  |  |  |
| Fast | 23 (18.0%) | 2 (3.6%) | -80% |
| Moderate | 82 (64.1%) | 36 (65.5%) | +2% |
| Slow | 23 (18.0%) | 17 (30.9%) | +72% |
| **Site of onset** |  |  |  |
| Bulbar | 23 (18.0%) | 9 (16.4%) | -9% |
| Spinal | 105 (82.0%) | 46 (83.6%) | +2% |
| **FVC (% predicted)** |  |  |  |
| $\geq 80$ | 84 (65.6%) | 38 (69.1%) | +5% |
| $< 80$ | 44 (34.4%) | 17 (30.9%) | -10% |
| **Age (years)** |  |  |  |
| $\geq 55$ | 67 (52.3%) | 24 (43.6%) | -17% |
| $< 55$ | 61 (47.7%) | 31 (56.4%) | +18% |

Long-term survival with masitinib was independent of the variables site of onset (bulbar or spinal), baseline respiratory function (categorized as above or below FVC of 80%), and patient age at randomization (categorized as above or below 55 years), as evidenced by the consistency of prevalence between the overall cohort and long-term survivor cohort, with long-term survivors being well-represented in all groups. Patients who achieved a long-term survival response under masitinib also included those with baseline predictive factors of moderate or severe loss of functionality and a moderate or slow ΔFS (defined as 1.1>ΔFS ≥0.3 points/month and ΔFS <0.3 points/month, respectively). In contrast, patients with baseline predictive factors indicating a complete loss of function (i.e., a score of 0 on at least one item of the ALSFRS-R scale) and fast-progressing disease (ΔFS ≥1.1 points/month) appeared to have a reduced likelihood of response (Table 3).

## Discussion

In the cohort initially assigned to receive masitinib at a dosage of 4.5 mg/kg/day in study AB10015, the 5-year survival rate was 42.3% from the onset of symptoms and 36.2% from the time of diagnosis. Notably, approximately half of these long-term survivors maintained a good QoL, as evidenced by the surrogate measure of a delayed need for mechanical assistance, such as permanent ventilation, gastrostomy, tracheostomy, or wheelchair dependence. Survival rates improved further among patients with a baseline ΔFS of less than 1.1 points/month, the primary efficacy analysis population of study AB10015, reaching 53% from the onset of symptoms when treatment was initiated early in the course of the disease.

Masitinib’s 5-year survival rate significantly surpasses historical benchmarks, which range from 7% to 27.8% (weighted average ∼23.5%), and compares favorably with riluzole-treated and untreated cohorts from large databases (riluzole-treated: ∼24%, untreated: ∼15% to 17%, rates from time of diagnosis) [Thakore 2020]. Currently, no established 5-year survival rate has been reported in peer-reviewed clinical studies or real-world data for patients treated with edaravone, AMX0035, or CNM-Au8. Real-world data on edaravone show lower survival rates at 3 years (<50% from onset) [Brooks 2022] than those of masitinib 4.5 mg/kg/day (73.1%). AMX0035 showed 1-year and 2-year survival rates of 80.9% and 51.6%, respectively, based on data from the CENTAUR trial involving 89 ALS patients treated with this therapy [Paganoni 2021], whereas masitinib 4.5 mg/kg/day had rates of 96.9% and 89.2%, respectively, at the same timepoints.

Overall, prolonged survival was achieved under this treatment regimen across a diverse sample of the study population, including patients with moderate or severe ALS severity, slow or moderate progression, and bulbar or spinal onset (Table 3). However, a reduced response prevalence was noted in patients with any complete loss of function or fast-progressing disease at baseline. This latter observation aligns with the main findings of study AB10015, which indicated that masitinib was less effective in patients with ΔFS ≥1.1 points/month [Mora 2020], and analysis showing that the subgroup ‘ALS prior to any complete loss of functionality’ experienced consistently greater benefits in terms of function, survival, and quality of life [Ludolph 2025]. Furthermore, it is consistent with masitinib’s mechanism of action, which has been shown to reduce the rate of motor neuron death by modulating mast cell and microglial activity, rather than regenerating motor neurons [Kovacs 2021, Trias 2020, Harrison 2020, Trias 2018, Trias 2017, Trias 2016]. In essence, masitinib is designed to slow disease progression rather than restore lost functions; therefore, administering masitinib as early as possible, while there is still good functionality, aligns better with its intended mechanism.

Currently, there is no established definition for a ‘long-term ALS survivor,’ with the literature suggesting a range of 5 to 10 years [Pupillo 2025; Pal 2024; Nona 2022; Spencer 2020; Westeneng 2018; Mateen 2010]. In defining long-term ALS survivors for this analysis, we considered the median survival from symptom onset to death based on recently published large cohort and population-based studies. For instance, a study conducted at the Mayo Clinic in Florida, involving 1,442 ALS patients from 2003 to 2019, found a median survival of 30 months, with an interquartile range (IQR) of 20 to 48 months [Engelberg-Cook 2024]. In a prospective cohort study using a German single-center ALS register from 2007 to 2019, patients who did not receive ventilation therapy during the course of the disease (n = 1,238) had a median survival from onset of 33.6 months (95% CI [31.6–35.7]), while those who received noninvasive ventilation (NIV) without subsequent tracheostomy invasive ventilation (n = 358) had a median survival from onset of 40.8 months (95% CI [37.2–44.3]) [Spittel 2021]. A retrospective Italian study involving 2,648 ALS patients from 2009 to 2013 found that the median survival time from onset to death/tracheostomy was 44 months (95% CI [42–46]) [Calvo 2017]. Similarly, in a retrospective observational study using the population-based ALS register in Lombardy, Italy, which included 828 patients from 1998 to 2012, the median survival for the entire cohort was 2.2 years, with an IQR of 1.1 to 4.4 years [Pupillo 2025]. Lastly, a study of 1,245 ALS patients from the Piemonte Register, Italy, during 2007–2016 found a median survival time of 2.67 years, with an IQR of 1.67 to 5.25 years for the entire cohort [Chio 2023]. Therefore, it is evident from the literature that surviving at least 5 years (60 months) from the onset of symptoms surpasses expectations regarding median survival and its associated upper quartile boundary, making it a suitable threshold for defining long-term ALS survivors.

The main strength of the 5-year survival analysis is that it was based on the masitinib 4.5 mg/kg/day arm of the intent-to-treat population with a long follow-up period. Considering that 9 of the 55 long-term survivors (16%) reached the 9-year survival threshold at the cutoff date (Table 2) and that the majority of these were still alive without the need for mechanical assistance, it will be of interest to revisit these data in the future to assess the 10-year survival rate. This analysis also has limitations that need to be acknowledged. First, it was a post-hoc evaluation that, in the absence of long-term placebo follow-up, relied on historical benchmark data as an indirect comparator. This approach inherently has constraints compared to the gold standard of randomized placebo-controlled data. Second, some data required for the ENCALS survival prediction model, such as genetic status (C9orf72 mutation), were unavailable and thus assumed to be negative, potentially impacting the accuracy of the predicted survival estimates. However, the original sensitivity analyses of the model showed that, even without this information, the model still provided accurate predictions [Westeneng 2018]. Additionally, the ENCALS model predicts a composite survival outcome that includes noninvasive ventilation >23 h/day and tracheostomy events, whereas the observed overall survival in this study was defined solely as the time from symptom onset to death. These differences in outcome measures may influence the interpretation of residual median survival benefits.

## Conclusion

The analysis of long-term survivors treated with masitinib in study AB10015 seemingly revealed a substantial survival advantage, as evidenced by the 5-year survival rate, when compared with historical benchmark data, and the residual median survival gain when compared with the ENCALS model. This finding corroborates reports from sites involved in the early access program for masitinib of remarkable longevity, including patients still receiving masitinib with survival of greater than 10 years and no mechanical assistance in the form of permanent ventilation, gastrostomy, tracheostomy, or wheelchair dependence. Overall, these results underscore masitinib’s potential to extend survival in a heterogeneous ALS population, particularly when administered before any complete loss of function and in a selected group excluding patients with rapidly progressing disease. Notably, the observation that long-term survivors are common across various ALS prognostic factors, such as patients with both slow or moderate baseline ΔFS, severe or moderate baseline functional severity, and bulbar or spinal onset, suggests the presence of a subpopulation whose disease progression is primarily driven by microglial and/or mast cell activity. If this is the case, then the recent discovery of a plasma biomarker that detects masitinib’s effect on pathological pro-inflammatory microglia could facilitate the early identification of those ALS patients most likely to achieve long-term survival [AB Science 2026]. Further prospective studies are needed to confirm these benefits and refine the patient selection criteria, ideally through the use of an easily accessible biomarker to optimize treatment outcomes.

## Data Availability

All data produced in the present study are available upon reasonable request to the authors

## FUNDING

This work and the associated publication costs were supported by AB Science (France).

## COMPETING INTERESTS

AM, CDM, and OH are employees and shareholders of AB Science. The remaining authors declare no conflicts of interest.

## CONTRIBUTORS

Conceptualization: ACL, AM, JSM, THP, JMC, GR, NBM, OH, PV; Data interpretation: AM; Writing — original draft preparation: CDM; Writing — draft review and editing: ACL, CDM, AM, JSM, THP, JMC, GR, NBM, OH, PV. All authors have reviewed the manuscript and approved the final version for submission.

## DISCLAIMER

Masitinib is under clinical development by the study funder AB Science. The sponsor was involved in the study design, interpretation of the data, writing of the report, and decision to submit the paper for publication.

## Notes

### Clinical Trial

NCT02588677

### Author Declarations

Study AB10015 was a multinational study. The protocol for study AB10015 received ethical approval from each of the 34 participating clinical sites. Provided below is a detailed list of the Institutional Review Boards / Ethics Committees of active sites (i.e. at least one patient randomized) that gave ethical approval for study AB10015. Local Ethics Committee: Secretaria Tecnica CEIC Hospital La Paz. Paseo De La Castellana, 261 Planta 8a Hospital General, 28046 Madrid, Spain. Local Ethics Committee: CAEI, Comite Autonomico De Ensayos Clinicos De Galicia. Edificio Administrativo San Lazaro S/N 15703 Santiago De Compostela La Coruna 1570.3 Santiago De Compostela, Spain. Local Ethics Committee: Montserrat Fernandez Recio Unitat De Suport Al CEIC HUVH (SCEI). Fundacio Hospital Universitari Vall D'hebron - Institut De Recerca (VHIR). Edifici 08035 Barcelona, Spain. Local Ethics Committee: Secretaria CEIC Hospital Universitari De Bellvitge Edifici Unitat De Recerca Feixa Llarga, S/N 08907 Hospitalet De Llobregat, Spain. Local Ethics Committee: IMIM-Institut De Recerca Hospital Del Mar Parc De Recerca Biomedica De Barcelona C/ Doctor Aiguader, 88 08003 Barcelona, Spain. Local Ethics Committee: Comite De Etica En Investigacion Clinica (CEIC) Larrea 1381 3o A C1117ABK, Ciudad Autonoma De Buenos Aires, Argentina. Local Ethics Committee: CIE Investigacion Biomedica, Fundacion Neurologica, Uruguay 840 C1015ABR, Ciudad Autonoma De Buenos Aires, Argentina. Local Ethics Committee: Comite De Etica En Investigacion HGA JM R. Mejia, General Urquiza 609, C1221ADC, Ciudad Autonoma De Buenos Aires, Argentina. Local Ethics Committee: Comite De Etica En Investigaciones Biomedicas, Montaneses 2325, C1428AQK, Ciudad Autonoma De Buenos Aires, Argentina. Local Ethics Committee: Comite De Etica En Investigacion Biomedica, Leandro N. Alem 1416, 2000 Rosario, Argentina. Local Ethics Committee: CIEM-Noa, Las Piedras 496, 4to Piso, 4000 Tucuman, Argentina. Local Ethics Committee: Neurological Clinic Gorkeho 1,811 01 Bratislava, Slovakia. Local Ethics Committee: Faculty Hospital Trnava, A. Zarnova 11, 917 75 Trnava, Slovakia. Local Ethics Committee: General Hospital With Policlinic Levoca Probstnerova Cesta, 2/3082, 054 01 Levoca, Slovakia. Local Ethics Committee: Hospital With Policlinic Stefana Kukuru Michalovce, Spitalska 2, 071 01 Michalovce, Slovakia. Local Ethics Committee: University Hospital Martin Kollarova 2, 036 59 Martin, Slovakia. Local Ethics Committee: CEIC Parque De Saude De Lisboa, Parque Saude De Lisboa Av. Do Brasil 53 Pav 17-A, 1749-004 Lisboa, Portugal. Local Ethics Committee: Comite De Etica En Investigacion Winsett Rethman, Capitan Lorenzo Aguilar # 669 Col. Obispado, 64060 Monterrey, Nuevo Leon, Mexico. Local Ethics Committee: Comitato Etico Interaziendale, AOU Citta Della Salute E Della Scienza Di Torino-Ospedale Le, Molinette Corso Bramante 88/90, 10126 Torino, Italy. Local Ethics Committee: Comitato Etico Milano Area C, Asst Grande Ospedale Metropolitano Niguarda Piazza, Ospedale Maggiore,3 20162 Milano, Italy. Local Ethics Committee: Comitato Etico Ospedale San Raffaele, Via Olgettina 60, 20132 Milano, Italy. Local Ethics Committee: Fondazione Policlinico Universitario A. Gemelli Un, Ce Fondazione Policlinico Universitario A. Gemelli Universita, Cattolica Del S. Cuore - Largo Agostino Gemelli, 8 00168 Roma, Italy. Local Ethics Committee: Comitato Etico Area Vasta Sud Est, A.O.U. Senese Policlinico Alle Scotte- Presso Farmacia, Ospedaliera,Viale Bracci 16, 53100 Siena, Italy. Local Ethics Committee: Comitato Etico Regione Liguria, C/O Irccs-Azienda Ospedaliera Universitaria San Martino-Ist, Largo Rosanna Benzi 10, 16132 Genova, Italy. Local Ethics Committee: Comitato Etico Provinciale Di Modena, Via Del Pozzo 71, 41124 Modena, Italy. Local Ethics Committee: Comitato Etico Istituto Auxologico Italiano, Via L. Ariosto 13, 20145 Milano, Italy. Local Ethics Committee: Ce Interaziendale Della Provincia Di Messina, A.O. Universitaria Policlinico "G. Martino" Via Consolare Valeria, 198125 Gazzi- Messina, Italy. Local Ethics Committee/ Scientific Council Of General Hospital Of Athens ''Evangelismos'', 45 Ipsilantou Str., 10676, Athens, Greece. Local Ethics Committee: 36 Areos Street, Paliao Faliro, 17562 Athens, Greece. Local Ethics Committee: University Medical Center (UMC) Utrecht Huispostnummer D01.343 Heidelberglaan 100, 3508 Ga Utrecht, Netherlands. Local Ethics Committee: MUHC Research Ethics Board (Neuroscience & Psychiatry), 3801 University Street, Room 686, H3A 2B4 Montreal, QC, Canada. Local Ethics Committee: Research Ethics Office, Room C819, 2075 Bayview Ave., M4N 3M5 Toronto, ON, Canada. Local Ethics Committee: Western University, Research, Support Services Building, Room 5150, N6G 1G9 London, ON, Canada. Institutional Review Board (IRB) Services, 372 Hollandview Trail, Suite 300, L4G 0A5 Aurora, ON, Canada.

### Summary of Updates

An author was added to the author list. Competing interests and author contribution information was updated accordingly.

